# Applying a Random Forest Approach in Predicting Health Status in Patients with Carotid Artery Stenosis 30 Days Post-Stenting

**DOI:** 10.1101/2024.08.14.24312025

**Authors:** Omar Qureshi, Carlos Mena-Hurtado, Gaëlle Romain, Jacob Cleman, Santiago Callegari, Kim G. Smolderen

**Affiliations:** Vascular Medicine Outcomes (VAMOS) Program, Department of Internal Medicine, Section of Cardiovascular medicine, Yale University, New Haven, CT; Yale School of Medicine, New Haven, CT; Department of Psychiatry, Section of Psychology, Yale University, New Haven, CT

**Keywords:** carotid artery stenosis, carotid artery stenting, health status, random forest

## Abstract

**Background:** Approximately 20% of ischemic strokes in the U.S. result from carotid artery stenosis. Carotid artery stenting (CAS) can reduce stroke risk, but variability in post-stenting health outcomes and their predictors are poorly understood. We examined 30-day post-CAS health status and derived its most important clinical predictors.

**Methods:** The SAPPHIRE Worldwide Registry measured pre-procedural and 30-day health status for carotid artery stenosis patients undergoing transfemoral-CAS using the SF-36 or EQ-5D between 2010-2014. Four health status scores were calculated: SF-36 Mental Component Summary (MCS) and Physical Component Summary (PCS), EQ-5D Index Value and Visual Analogue Scale (VAS). Random Forest models ranked 66 pre-procedural candidate variables by relative importance in predicting 30-day post-CAS health status, stratified by symptomatic status. Variables with highest importance were used to develop predictive multivariable linear regression models. Model accuracy was assessed via Out of Bag accuracy and R-square, respectively.

**Results:** Health status was assessed using the SF-36 in 3,017 patients and EQ-5D in 3,390 patients. Random Forest models demonstrated high accuracy (86.7% - 95.2%) and identified nine key predictors of post-stenting health status: pre-procedural health status (RI 100%), Modified Rankin Scale (RI 26.2-76.5%), NIH Stroke Scale (RI 12.1-28.0%), history of stroke (RI 9.2- 19.8%), congestive heart failure (RI 12.3-19.7%), spinal immobility (RI 6.7-31.0%), diabetes mellitus (RI 8.1-32.9%), severe pulmonary disease/COPD (RI 13.8-45.6%), and non-Hispanic/Latino ethnicity (RI 8.4-32.4%). Multivariable linear regression models explained ∼36- 61% of health status variance, with the asymptomatic SF-36 PCS model explaining 61%. SF-36 PCS and MCS models outperformed EQ-5D Index Value and VAS models regarding R-square and visual fit of observed vs. predicted values.

**Conclusions:** We successfully derived prediction models for patient-centered outcomes following CAS which partially explained 30-day post-CAS health status outcomes. Pre-procedural health status, stroke scale scores, and medical comorbidities should be considered when discussing health status benefits in pre-CAS treatment decision discussions.

## INTRODUCTION

In the United States, around 20% of ischemic strokes present secondary to carotid artery stenosis.^1^ Carotid revascularization in carotid artery stenosis patients can decrease the risk of ischemic stroke, which can be as high as 11% in symptomatic patients.^1,2^ Revascularization can be achieved through several means, including carotid artery stenting (CAS) or carotid endarterectomy (CEA). CAS has been shown to be non-inferior to CEA in severe carotid artery stenosis patients.^3^ However, revascularization procedures confer non-negligible risk of post-procedural sequelae such as periprocedural stroke and mortality, which occurred in ≤5.6% and ≤1.4% of Vascular Quality Initiative patients, respectively, undergoing urgent revascularization.^2,4^ Therefore, revascularization benefits must be weighed against peri- procedural risks which may cause patients’ health to not improve or worsen post-revascularization. With this in mind, the Centers for Medicare & Medicaid Services (CMS) advises that, in addition to medical therapy, CAS is reasonable to consider in symptomatic patients with ≥50% stenosis or asymptomatic patients with ≥70% stenosis.^5^

Besides stroke risk, CAS has implications for individuals’ health status. Increasingly, health status measures are incorporated in clinical trial endpoints and benchmarks for routine clinical care evaluation. While short-term health status changes have been noted for CAS over CEA at 2 weeks post-intervention, it is unknown what the main predictors of post-stenting health status outcomes are in carotid artery stenosis patients.^6^ As current trials actively compare medical management, CAS, and CEA head-to-head in managing carotid artery stenosis, it is important to compare health status outcomes from this clinical trial work against real-world cohorts. This comparison can serve as context for expected benchmarks and interpreting effect sizes for meaningful health status changes associated with treatment.^7^ Additionally, understanding which clinical patient profiles are associated with health status improvement following CAS is critical in informing shared decision-making in carotid artery stenosis.

We aimed to identify the most important patient characteristics in predicting carotid artery stenosis patients’ health status 30 days post-stenting using a Random Forest-based machine learning approach in the SAPPHIRE Worldwide Registry.

## METHODS

### Study Design and Patient Cohorts

The SAPPHIRE Worldwide Registry is an international (U.S., Canada), prospective, observational registry using data from 390 clinical sites encompassing patients enrolled between 2006 and 2014 who underwent either transfemoral-CAS or CEA for asymptomatic or symptomatic carotid artery stenosis. The registry’s design has been previously described.^8,9^ The 36-item Short Form Survey (SF-36) was administered to patients enrolled between 2010-2012 and the Euroqol-5 Dimension (EQ-5D) to patients enrolled between 2012-2014^10,11^. The NIH Stroke Scale and Modified Rankin Scale were administered to patients enrolled between 2010- 2014.^12,13^ This research was exempt from annual IRB review under section 45CFR46.104(4). Patient information was obtained from medical records and interviews conducted by clinicians and clinical volunteers.

From the SAPPHIRE Worldwide registry, we derived two patient cohorts: SF-36 and EQ- 5D. Inclusion criteria were: 1) stenting performed between 2006-2014; 2) stenting performed during the years in which the respective health measure for each cohort was assessed (i.e., 2010- 2012 for SF-36 and 2012-2014 for EQ-5D); 3) patient is ≥ 18 years old; 4) patient has ≥1 anatomic or physiological risk factor placing them at high risk for surgery.^8,9^ Exclusion criteria were: 1) Instructions for Use (IFU) outlined for the stent (Cordis PRECISE Nitinol Stent System) used in the SAPPHIRE study was not followed during stenting or information regarding whether IFU were followed is missing, 2) no stent was deployed during the procedure, 3) health status (i.e. SF-36 or EQ-5D) was not assessed at baseline or <28 days prior to procedure, 4) health status was not assessed 30±7 days post-stenting.

Symptomatic patients have symptoms resulting from their carotid artery stenosis or associated pathology (i.e., stroke, transient ischemic attack), such as changes in speech, sensation, movement, coordination, or behavior. Asymptomatic patients lack these symptoms. Symptomatic carotid artery stenosis patients face markedly higher stroke risk compared to asymptomatic patients regardless of degree of stenosis.^14^ Accordingly, asymptomatic and symptomatic patients are clinically evaluated and treated differently.^1,2,4,5,14^ We therefore stratified SF-36 and EQ-5D cohorts by symptomatic status to contrast findings across these two clinical profiles.

### Health Status Measures

The main outcome was patient health status scores at 30±7 days post-transfemoral-CAS on the following health status measures: SF-36 PCS, SF-36 MCS, EQ-5D Index Value, or EQ- 5D VAS.

The SF-36 is a standardized, 36-item generic health status instrument measuring physical and mental health status across eight domains: physical functioning, role-physical, bodily pain, general health, vitality, social functioning, role-emotional, and mental health.^11^ Scores across dimensions are used to calculate Physical Component (PCS) and Mental Component (MCS) Summary scores, with country-specific weights applied to each domain based on whether the PCS or MCS is being calculated. Each component is scored from 0-100. Higher scores indicate better health status. PCS and MCS scores can be converted into utility scores for health economic analyses and are used globally to evaluate cardiovascular disease patients.^15,16^

The EQ-5D is a standardized, generic health status instrument which asks patients to rate their health across five dimensions: mobility, self-care, usual activities, pain/discomfort, and anxiety/depression.^10^ Health across dimensions is used to calculate a utility score reflecting patients’ perceived positive and negative health effects post-intervention. Country-specific weights are applied to utility scores to generate the EQ-5D Index Value scores. A score <0 indicates health states perceived worse than death, 0 indicates health states like death, and 1 indicates full health. EQ-5D Visual Analogue Scale (VAS) scores use patients’ visual self-rating of health from 0-100.^10^ Higher scores indicate better health status. The EQ-5D’s brevity and ease of use make it one of the most frequently used instruments for calculating quality-adjusted life years in health economic analyses.^10,17–19^ Low EQ-5D scores correlate with level of anatomic disease, disease progression, and risk of poor clinical outcomes in cardiovascular disease patients.^17–19^

### Candidate Variables

We considered 66 pre-procedural candidate variables, listed in **Table 1**, including pre-procedural health status instrument scores. We combined variables for which prevalence was ≤1% among patients in any cohort within a “Rare Characteristics” variable in our analysis. Candidate variables were categorized into the following categories: demographic, high-risk criteria, medical history, neurological history, procedural and lesion characteristics, and baseline stroke scale or health status scores. High-risk criteria are defined as the most common risk factors among SAPPHIRE-eligible patients who underwent CEA.

**Table 1.** Baseline characteristics SAPPHIRE Worldwide registry patients and for whom health status was assessed using the EQ-5D or the SF-36. Values are presented as N (%), unless otherwise specified.

|  | **EQ-5D Cohort,**<br>**N=3,930** | **SF-36 Cohort,**<br>**N=3,017** | **P-Value** | ***d*** |
| --- | --- | --- | --- | --- |
| **Demographic Factors** |  |  |  |  |
| Age (Mean, SD) | 72.43 (9.26) | 72.00 (9.30) | 0.06 | 0.05 |
| Race |  |  | 0.25 | 0.04 |
| White | 3,636 (92.5) | 2,799 (92.8) |  |  |
| Black or African American | 192 (4.9) | 127 (4.2) |  |  |
| Other | 102 (2.6) | 91 (3.0) |  |  |
| Sex |  |  | 0.16 | 0.03 |
| Male | 2,483 (63.2) | 1,856 (61.5) |  |  |
| Female | 1,447 (36.8) | 1,161 (38.5) |  |  |
| Ethnicity |  |  | 0.71 | 0.01 |
| Hispanic Or Latino | 125 (3.2) | 100 (3.4) |  |  |
| Not Hispanic or Latino | 3,734 (96.8) | 2,841 (96.6) |  |  |
| Public Insurance | 3,216 (81.8) | 2,408 (79.8) | 0.03 | 0.05 |
| Private Insurance | 1,365 (34.7) | 1,149 (38.1) | <0.01 | 0.07 |
| **High-Risk Criteria** |  |  |  |  |
| Congestive Heart Failure | 575 (14.6) | 431 (14.3) | 0.69 | 0.01 |
| Unstable Angina | 131 (3.3) | 94 (3.1) | 0.61 | 0.01 |
| Severe Pulmonary Disease/COPD | 870 (22.1) | 702 (23.3) | 0.26 | 0.03 |
| Cardiac Arrhythmia | 609 (15.5) | 445 (14.7) | 0.39 | 0.02 |
| Abnormal Cardiac Stress Test | 502 (12.8) | 404 (13.4) | 0.45 | 0.02 |
| Age > 75yrs | 1,665 (42.4) | 1,255 (41.6) | 0.52 | 0.02 |
| ≥2 Proximal or Major Diseased Coronary Arteries With >70% Stenosis | 182 (4.6) | 150 (5.0) | 0.51 | 0.02 |
| Spinal Immobility | 69 (1.8) | 51 (1.7) | 0.84 | 0.01 |
| Bilateral Artery Stenosis | 401 (10.2) | 316 (10.5) | 0.71 | 0.01 |
| **Medical History** |  |  |  |  |
| Body Mass Index (kg/m2)<br>(Mean, SD) | 28.39 (5.73) | 28.26 (5.70) | 0.33 | 0.02 |
| Coronary Artery Disease | 2,196 (55.9) | 1,633 (54.1) | 0.15 | 0.04 |
| First-degree relative with<br>Coronary Artery Disease | 341 (8.7) | 231 (7.7) | 0.13 | 0.04 |
| Diabetes Mellitus | 1,442 (36.7) | 1,046 (34.7) | 0.08 | 0.13 |
| Hyperlipidemia | 3,216 (81.8) | 2,312 (76.6) | < 0.01 | 0.04 |
| Hypertension | 3,351 (85.3) | 2,531 (83.9) | 0.12 | 0.03 |
| Renal Failure/ Insufficiency | 186 (4.7) | 122 (4.0) | 0.17 | 0.02 |
| Severe Aortic/Mitral Valve<br>Disease | 132 (3.4) | 92 (3.0) | 0.47 | 0.06 |
| Synchronous, Severe<br>Cardiac and Carotid Disease | 104 (2.6) | 110 (3.6) | 0.02 | 0.02 |
| Aortic Valve Replacement | 88 (2.2) | 77 (2.6) | 0.40 | 0.03 |
| Coronary Artery Bypass<br>Graft | 1,029 (26.2) | 752 (24.9) | 0.23 | 0.08 |
| Coronary Percutaneous<br>Revascularization | 890 (22.6) | 586 (19.4) | <0.01 | 0.01 |
| History of Smoking | 2,100 (53.4) | 1,596 (52.9) | 0.66 | 0.04 |
| Myocardial Infarction | 696 (17.7) | 581 (19.3) | 0.10 | 0.01 |
| Post Radiation Treatment | 241 (6.1) | 180 (6.0) | 0.77 | 0.09 |
| **Neurological History** |  |  |  |  |
| History of Stroke | 758 (19.4) | 687 (22.9) | < 0.01 | 0.02 |
| History of TIA | 782 (20.0) | 619 (20.6) | 0.54 | 0.09 |
| No Neurological History | 1,919 (49.1) | 1,337 (44.6) | < 0.01 | 0.00 |
| New neurological deficit<br>when the patient left the<br>angiography suite | 67 (1.7) | 51 (1.7) | 0.96 | 0.02 |
| Prior Stent or Carotid PTA | 241 (6.1) | 200 (6.6) | 0.40 | 0.02 |
| **Procedural and Lesion Characteristics** |  |  |  |  |
| Side of Treated Lesion |  |  | 0.32 | 0.00 |
| Right | 1,934 (49.2) | 1,521 (50.4) |  |  |
| Left | 1,996 (50.8) | 1,496 (49.6) |  |  |
| Additional PCI planned for<br>contralateral lesion | 219 (5.6) | 166 (5.5) | 0.90 | 0.04 |
| Contralateral Carotid<br>Occlusion | 360 (9.2) | 310 (10.3) | 0.12 | 0.04 |
| Arch Type |  |  | 0.34 | 0.04 |
| I | 1,797 (45.7) | 1,403 (46.5) |  |  |
| II | 1,451 (36.9) | 1,067 (35.4) |  |  |
| III | 496 (12.6) | 381 (12.6) |  |  |
| Not Done | 186 (4.7) | 166 (5.5) |  |  |
| Imaging modality used during patient evaluation |  |  |  | 0.13 |
| MRA Angiography | 145 (3.7) | 196 (6.5) | < 0.01 | 0.13 |
| Traditional Angiography | 3,748 (95.4) | 2,784 (92.3) | < 0.01 | 0.09 |
| Ultrasound | 903 (23.0) | 808 (26.8) | < 0.01 | 0.03 |
| Other imaging modality used | 275 (7.0) | 190 (6.3) | 0.25 | 0.09 |
| Reference diameter (mm) (Mean, SD) | 6.01 (1.32) | 5.89 (1.34) | < 0.01 | 0.00 |
| Target Lesion % Diameter Stenosis (Mean, SD) | 85.15 (8.85) | 85.18 (8.95) | 0.90 | 0.06 |
| Target Lesion Length (mm) (Mean, SD) | 18.38 (8.66) | 17.85 (8.83) | 0.01 | 0.01 |
| Total length of stented segment (mm) (Mean, SD) | 34.99 (7.70) | 34.92 (7.69) | 0.73 | 0.01 |
| Geometry - Eccentric Target Lesion | 2,238 (56.9) | 1,702 (56.4) | 0.66 | 0.05 |
| Geometry - Thrombus Present Within Target Lesion | 131 (3.3) | 76 (2.5) | 0.05 | 0.04 |
| Geometry - Ulcerated Target Lesion | 1,069 (27.2) | 878 (29.1) | 0.08 | 0.00 |
| Geometry - Other | 1,246 (31.7) | 951 (31.5) | 0.87 | 0.03 |
| High Cervical Internal Carotid Artery/ Common Carotid Artery Lesions Below the Clavicle | 432 (11.0) | 304 (10.1) | 0.22 | 0.08 |
| Severe Tandem Lesions | 71 (1.8) | 93 (3.1) | < 0.01 | 0.12 |
| Quantitative Lesion Calcification (Visual) |  |  | < 0.01 | 0.12 |
| None | 920 (23.6) | 859 (29.0) |  |  |
| Mild | 1,370 (35.2) | 986 (33.2) |  |  |
| Moderate | 1,091 (28.0) | 771 (26.0) |  |  |
| Severe | 516 (13.2) | 350 (11.8) |  | 0.10 |
| Concentric Lesion Calcification | 1,251 (32.1) | 867 (29.2) | 0.01 | 0.06 |
| Circumferential Lesion Calcification | 616 (15.8) | 437 (14.7) | 0.22 | 0.03 |
| Other Qualitative Lesion Calcification | 1,871 (47.7) | 1,584 (52.6) | < 0.01 | 0.06 |
| Target lesion ≥ 3mm Width | 533 (13.7) | 346 (11.7) | 0.01 | -0.04 |
| **Baseline Stroke Scale or Health Status Scores** |  |  |  |  |
| Modified Rankin Scale Score (Mean, SD) | 1.41 (0.91) | 1.45 (0.93) | 0.08 | -0.05 |
| Stroke NIH Scale Score (Mean, SD) | 0.52 (1.58) | 0.59 (1.58) | 0.06 | 0.05 |
| EQ-5D Index Value (Mean, SD) | 0.83 (0.18) | N/A | N/A | N/A |
| EQ-5D VAS (Mean, SD) | 69.27 (22.04) | N/A | N/A | N/A |
| SF-36 PCS (Mean, SD) | N/A | 39.07 (10.78) | N/A | N/A |
| SF-36 MCS (Mean, SD) | N/A | 48.38 (9.45) | N/A | N/A |

Abbreviations: *d* = standard difference with absolute value $\geq 0.2$ indicating medium to large effect size, SD = Standard Deviation; VAS = Visual Analogue Scale, PCS = Physical Component Summary, MCS = Mental Component Summary, COPD = Chronic Obstructive Pulmonary Disease, TIA = Transient Ischemic Attack, PTA = Percutaneous Transluminal Angioplasty, PCI = Percutaneous Coronary Intervention, NIH = National Institutes of Health

Pre-procedural NIH Stroke Scale and Modified Rankin Scale scores were included as candidate variables. The NIH Stroke Scale and Modified Rankin Scale measure patient disability in the clinical evaluation of stroke patients and in stroke-related clinical trials.^12,13^ The NIH Stroke Scale is a 15-item assessment of stroke-related neurological deficits administered to acute stroke patients.^12,20^ The Modified Rankin Scale is a 6-point measure of global disability measured by patients’ functional ability and independence in completing activities of daily living.^13,21^ Both correlate strongly with stroke patient health status post-intervention, can be reliably reproduced, and predict inpatient length of stay for stroke patients.^21–23^

### Statistical Analyses

Patient characteristics were described and compared between the SF-36 and EQ-5D cohorts. Additional comparisons were made between symptomatic and asymptomatic patients within each cohort.

Continuous variables were summarized as means with standard deviations and compared using Student t-tests. Categorical variables were summarized as frequencies and were compared using Chi-square or Fisher’s exact tests as appropriate. We assessed effect sizes between cohorts using standard differences (*d*). An absolute value |*d*| ≤0.1, ≤0.2, ≤0.5, or ≤0.8 was considered negligeable, small, moderate, or large effect size, respectively.^24^

### Random Forest Model development

We constructed eight Random Forest (RF) models: one for each health status scores (SF- 36 PCS and MCS, EQ-5D Index and VAS) stratified by patient symptomatic status (symptomatic or asymptomatic). Each RF ranked 66 candidate variables based on their relative importance in predicting patient health status 30 days post-stenting.

A Random Forest (RF) is a machine learning algorithm which constructs multiple decision trees to rank variables by relative importance in predicting an outcome. RF was selected for its superior performance in handling variables with multicollinearity and non-linear effects on the outcome.^25,26^ To reduce overfitting and multicollinearity, RFs train each tree using subset of randomly selected variables using bootstrap sampling (constructed using 2/3 of the data). The remaining 1/3, known as Out of Bag (OOB), was used to assess RF error.^25^

First, SF-36 and EQ-5D sub-cohorts stratified by symptomatic status were randomly divided into training samples (80% of data) to build RFs and testing samples (20%) to validate RFs. We assessed candidate variable similarity between these samples using standard differences (*d*).

Second, to improve RF performance and ensure that the RFs will be generalizable to new data, we tuned the RF hyperparameters, the number of decision trees and the number of randomly selected variables, using a grid search method in the training sample. To minimize the RFs’ OOB error, RFs were then trained using 500 decision trees, with each tree considering a random subset of 30 out 66 candidate variables.

Then, using testing sample data, 66 candidate variables were ranked based on their relative importance in predicting 30-day post-stenting health status scores. Relative importance was calculated based on prediction error attributable to each variable using Breiman-Cutler importance.^25^ For each RF, we determined the most important variables via graphical relative importance plots as this represents an intuitive interpretation of variable importance.

Finally, using testing sample data, we evaluated RF accuracy based on the OOB accuracy (i.e., -1*[OOB error - maximum score on health status measure]), which estimates RF generalization to unseen data, and the root mean square error (RMSE) which indicates the average difference between the individual predictions and observed values.^24^

### Multivariable Linear Regression Model Development

Since RFs do not assess the directionality of a variable’s effect, we used linear regression models to create more easily implementable and interpretable clinical models. We constructed eight multivariable linear regression models in regards of the RFs previously constructed: one for each health status instrument stratified by patient symptomatic status. Each regression model included the variables previously identified as the “most important” in any RF with care center incorporated as a random effect, to derive coefficients (β) with 95% confidence interval (CI). Model accuracy was assessed based on RMSE and were compared with those of RFs. Additionally, model performance was evaluated based on R-square, which provide the percentage of variance in outcomes explained by the model, along with plots of observed versus predicted health status scores.

All statistical tests were two-tailed with an alpha of 0.05 as a criterion for statistical significance. All statistical analyses were performed using Stata Statistical Software: Release 17 (StataCorp LLC; College Station, TX).

### Missing Data

Missing data for candidate variables in RFs were handled during the RF’s growth process, as RFs consider a subset of randomly selected variables during each tree’s construction. Missing data for adjustment variables in linear regression models were handled using multiple imputation by chained equation, with 10 imputed data sets pooled using Rubin’s Rules.^24^

## RESULTS

### Overview of SF-36 and EQ-5D cohorts and rare characteristics

After inclusion and exclusion criteria, the SF-36 cohort consisted of 3,017 participants (14.36% of registry) and EQ-5D cohort consisted of 3,390 participants (18.71% of registry) (**Figures 1a-b**). After stratifying cohorts by symptomatic status, SF-36 sub-cohorts included 2,098 asymptomatic patients and 919 symptomatic patients. EQ-5D sub-cohorts included 2,840 asymptomatic patients and 1,090 symptomatic patients.

**Figure 1a.**
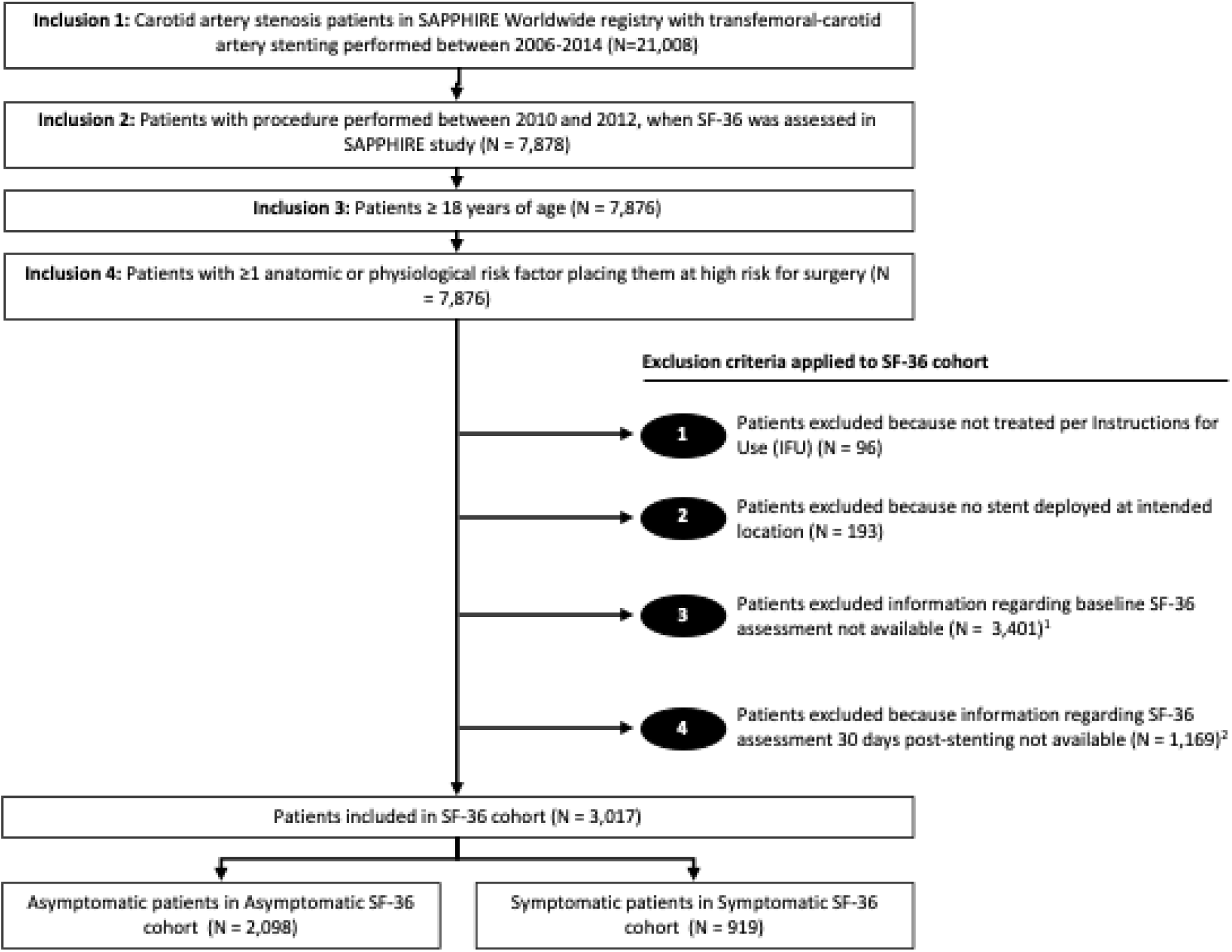
SF-36 cohort derivation from the SAPPHIRE registry.

**Figure 1b.**
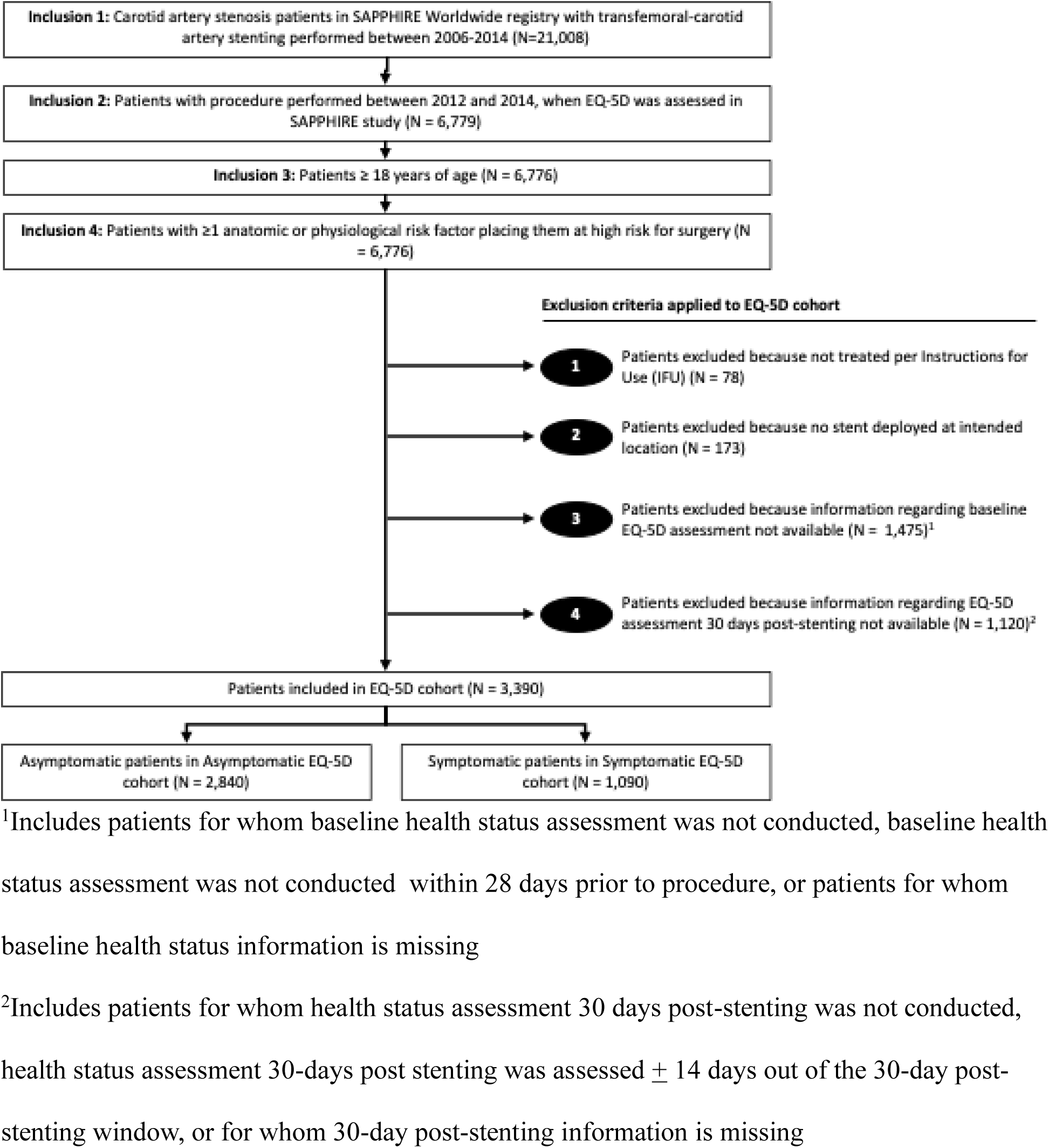
EQ-5D cohort derivation from the SAPPHIRE registry.

We categorized eleven variables as “Rare Characteristics” in RFs: having contralateral laryngeal palsy, being on or under evaluation for an organ transplant list, having a tracheostomy stoma, requiring cardiac valve surgery, having a recent myocardial infarction, having no health insurance, previous mitral valve replacement, previous extracranial-intracranial bypass, having significant aortic arch disease, having no medical history available, and history of open heart surgery.

The rate of missing data ranged from 0% to 4.3% (target lesion length variable) in the SF- 36 cohort and from 0% to 4.5% (lesion reference diameter variable) in the EQ-5D cohort.

### Patient characteristics for EQ-5D and SF-36 cohorts

Patient characteristics and candidate variables, including baseline health status scores, for the EQ-5D and SF-36 cohorts are described and compared in **Table 1**. Baseline characteristics across the SF-36 vs. EQ-5D cohorts were not different (all |*d*| < 0.20), with a mean age of 72.00±9.30 years vs. 72.43±9.26 years, 92.8% vs. 92.5% of patients being White, and 61.5% vs. 63.2% of patients being male, respectively. In the SF-36 cohort, 69.5% of patients were asymptomatic and 72.3% in the EQ-5D cohort (*d* = 0.06). Patient characteristics for SF-36 and EQ-5D cohorts stratified by symptomatic status are described and compared in **Supplemental Tables 1 and 2**, respectively. Across asymptomatic and symptomatic SF-36 and EQ-5D sub-cohorts, there were moderate-to-large effect sizes in the following variables: Modified Rankin Score, NIH Stroke Scale score, bilateral artery stenosis, coronary artery disease, history of stroke, history of transient ischemic attack, no neurological history, target lesion % diameter stenosis, ulcerated target lesion, and thrombus within target lesion (|*d*| ≥ 0.20).

### RFs: Relative importance in predicting 30 day post-stenting health status and performances

The relative importance and ranking of the 66 candidate variables in predicting 30-day post-stenting SF-36 scores (MCS and PCS) and EQ-5D (Index Value and VAS) stratified by symptoms status are in **Supplemental Tables 3 and 4**, respectively. Random forest plots of variables with highest relative importance for each health status outcome stratified by symptomatic status are in **Supplemental Figures 1-2.**

We identified nine pre-procedural variables as the “most important” variables in predicting the 30-day health status scores. Baseline health status score emerged as the most important variable, with a relative importance (RI) of 100% across all health status outcomes stratified by symptom status. The other “most important” variables were: Modified Rankin Scale score (RI 26.2-76.5%), NIH Stroke Scale score (RI 12.1-28.0%), history of stroke (RI 9.2-19.8%), congestive heart failure (CHF) (RI 12.3-19.7%), spinal immobility (RI 6.7-31.0%), diabetes mellitus (RI 8.1-32.9%), severe pulmonary disease/chronic obstructive pulmonary disease (COPD) (RI 13.8-45.6%), and not having Hispanic/Latino ethnicity (RI 8.4-32.4%).

The RFs showed good performance across the eight RFs in predicting post-stenting health status scores with OOB accuracy ranging from 86.70% to 95.16% (**Table 2**). RF RMSEs are in **Table 2**.

**Table 2.**
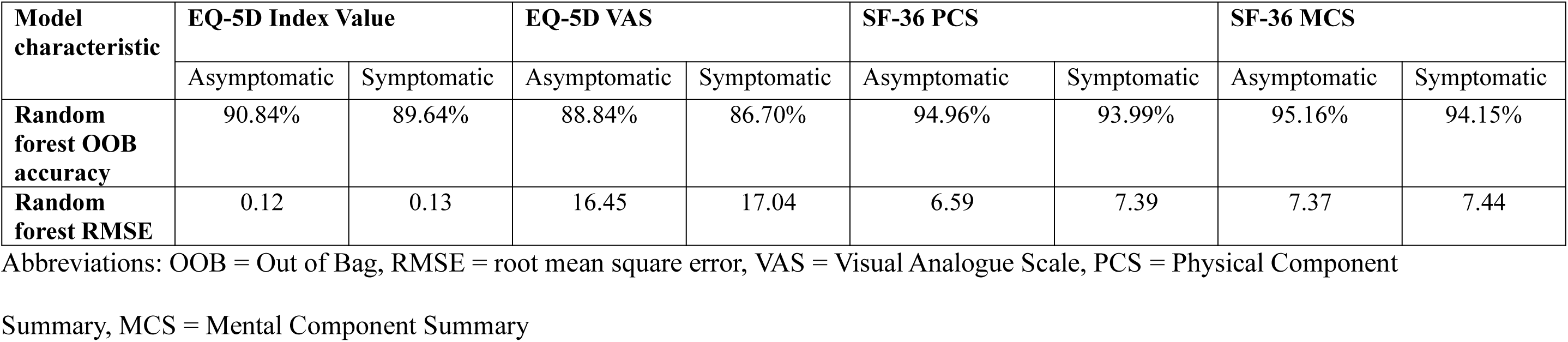
Random Forest model performance for each health status measure stratified by symptom status.

| Model characteristic | EQ-5D Index Value |  | EQ-5D VAS |  | SF-36 PCS |  | SF-36 MCS |  |
| --- | --- | --- | --- | --- | --- | --- | --- | --- |
|  | Asymptomatic | Symptomatic | Asymptomatic | Symptomatic | Asymptomatic | Symptomatic | Asymptomatic | Symptomatic |
| Random forest OOB accuracy | 90.84% | 89.64% | 88.84% | 86.70% | 94.96% | 93.99% | 95.16% | 94.15% |
| Random forest RMSE | 0.12 | 0.13 | 16.45 | 17.04 | 6.59 | 7.39 | 7.37 | 7.44 |

Abbreviations: OOB = Out of Bag, RMSE = root mean square error, VAS = Visual Analogue Scale, PCS = Physical Component
Summary, MCS = Mental Component Summary

### Multivariable linear regression

When evaluating 30-day SF-36 PCS and MCS, higher pre-procedural scores were associated with better 30-day health status regardless of patient symptomatic status (p-values <0.01). In asymptomatic patients, having severe pulmonary disease/COPD is highly associated with decreased 30-day SF-36 PCS only (β = -1.15, 95% CI -1.88; -0.41, p-value <0.01), while higher pre-procedural Modified Rankin Scale Score is highly associated with decreased 30-day SF-36 PCS and MCS (SF-36 PCS: β = -0.92, 95% CI -1.39; -0.44, p-value <0.01; SF-36 MCS: β = -0.79, 95% CI -1.27; -0.31, p-value <0.01) (**Figure 4**). In symptomatic patients, having diabetes mellitus was associated with decreased SF-36 PCS only (β = -1.23, 95% CI -2.30; -0.16, p-value = 0.02) (**Figure 5**).

Similarly, when evaluating 30-day EQ-5D Index Value and VAS, higher pre-procedural scores were associated with increase in 30-day health status regardless of patient symptomatic status (p-values <0.01). In asymptomatic patients, CHF and severe pulmonary disease/COPD were most strongly associated with decreased 30-day EQ-5D Index Value and VAS (**Supplemental Figure 3**), while in symptomatic patients, higher pre-procedural Modified Rankin Scale Score was most strongly associated with decreased 30-day EQ-5D Index Value and VAS (**Supplemental Figure 4**).

The performances of the eight multivariable linear regression models are summarized in **Figures 2-3**. When predicting health status in asymptomatic patients, R-squares ranged from 37% for the model predicting 30-day EQ-5D VAS scores in symptomatic patients to 61% for the model predicting SF-36 PCS scores. Moreover, the RMSEs of regression models were comparable to those of RFs, suggesting that including additional variables to predict the health status at 30 days post-stenting would not improve the models’ predictive performance.

**Figure 2.**
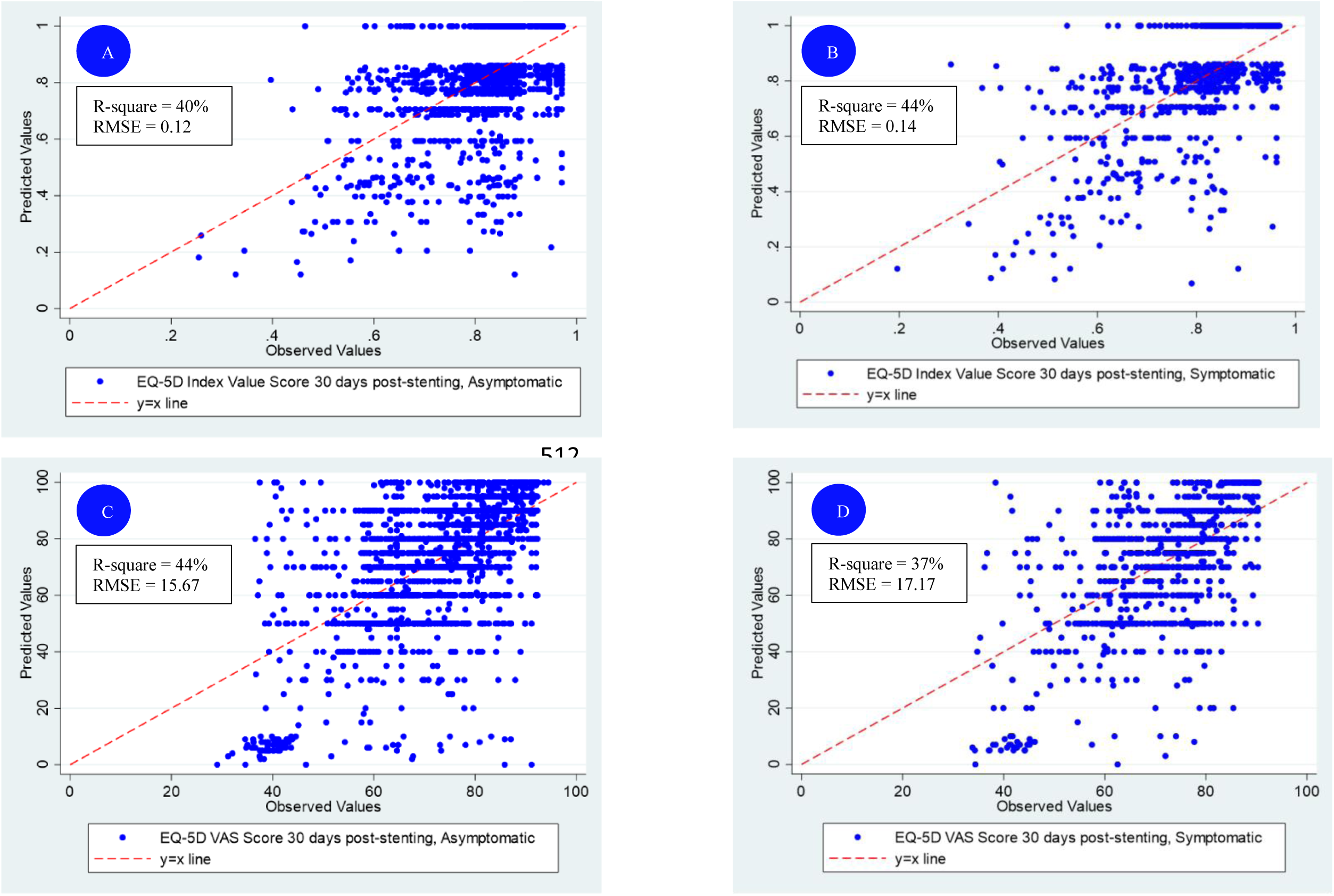
30 days post-stenting observed vs. predicted EQ-5D scores from multivariable linear regression models with health status measured using the EQ-5D Index Value in (A) asymptomatic and (B) symptomatic patients, and the Visual Analogue Scale (VAS) in in (C) asymptomatic and (D) symptomatic patients.

**Figure 3.**
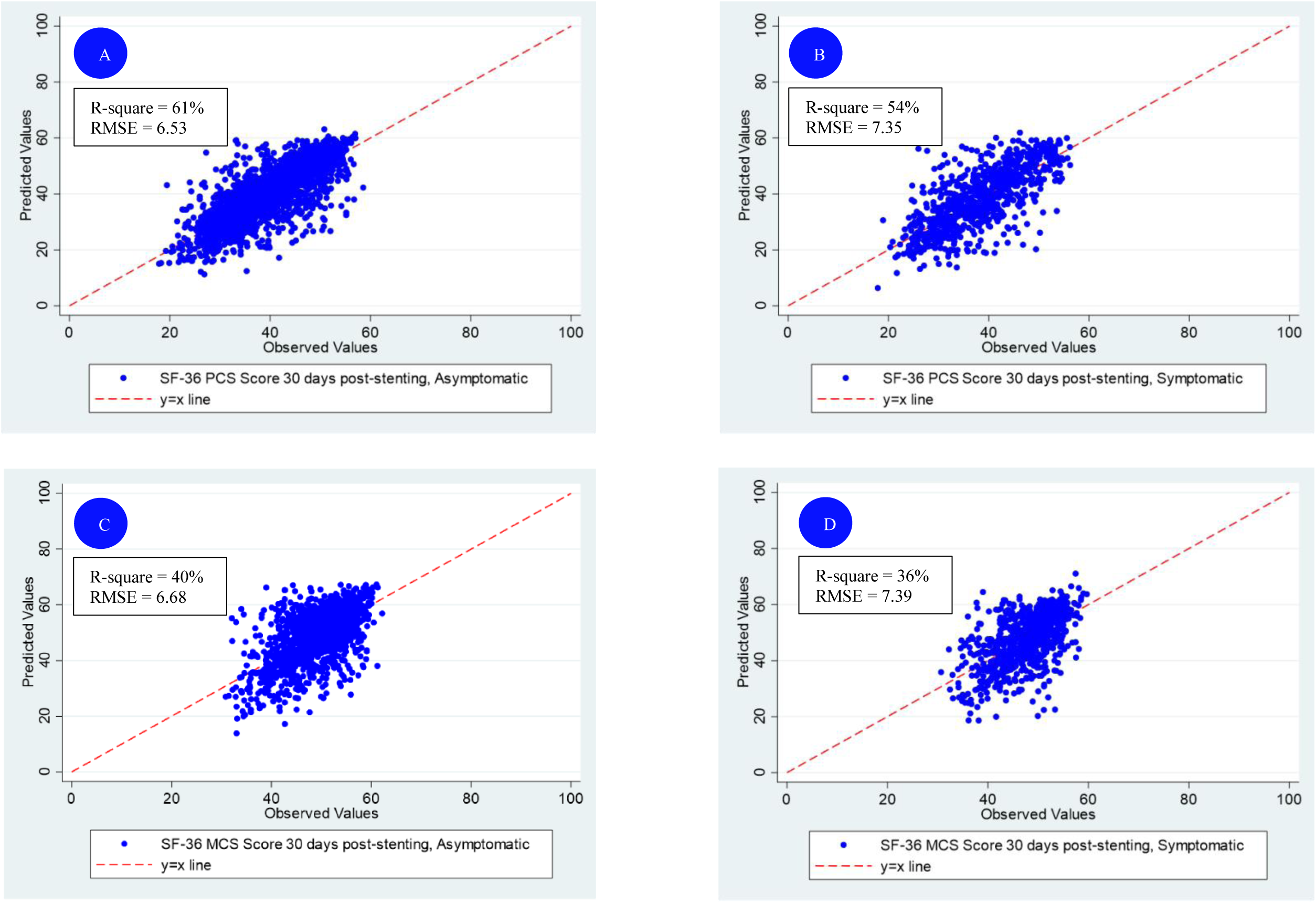
30 days post-stenting observed vs. predicted SF-36 scores from multivariable linear regression models with health status measured using the SF-36 Physical Component Summary (PCS) in (A) asymptomatic and (B) symptomatic patients, and the Mental Component Summary (MCS), in (C) asymptomatic and (D) symptomatic patients.

**Figure 4.**
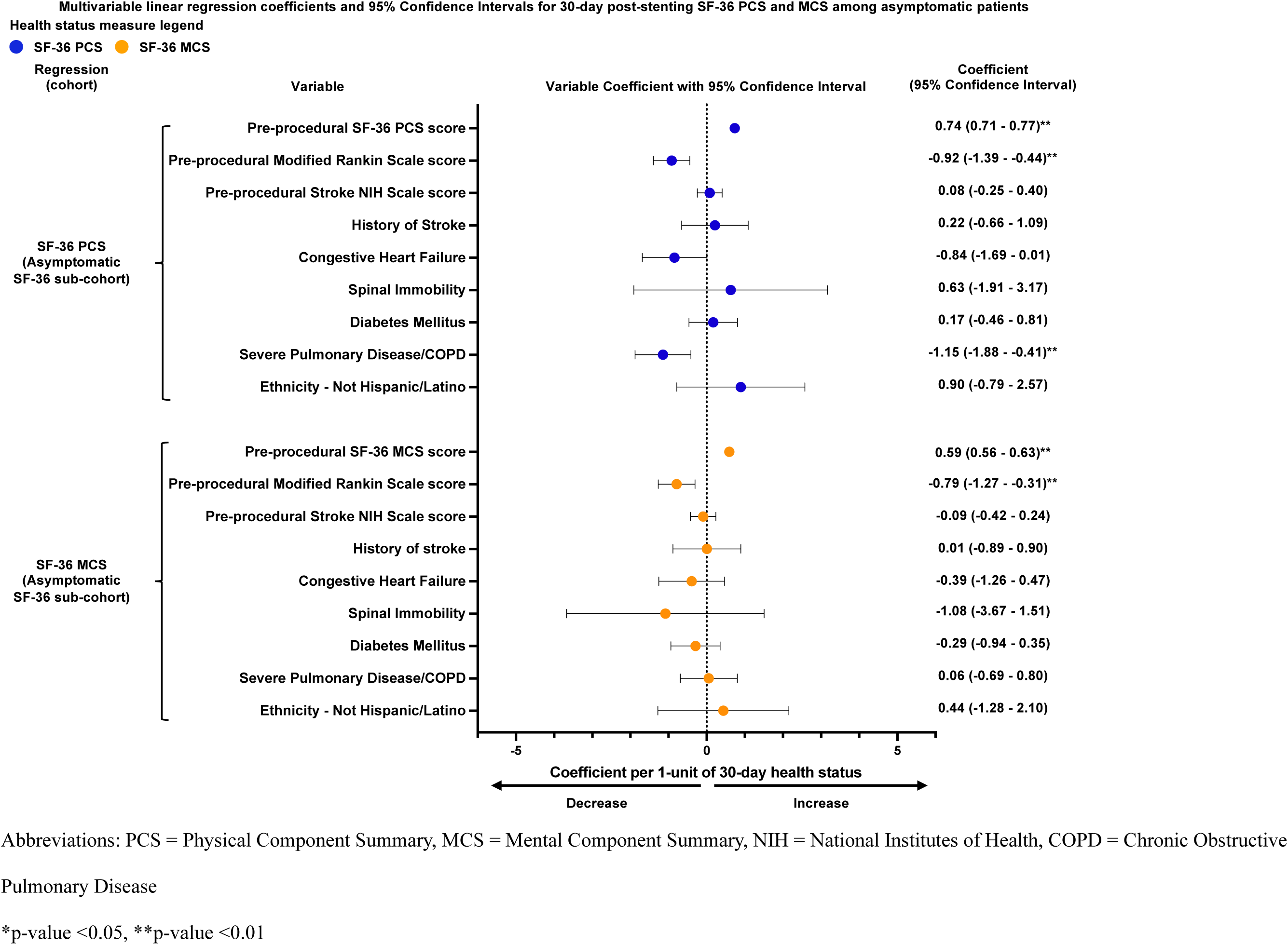
Coefficients and 95% Confidence intervals derived from multivariable linear regression models to predict 30 day post- stenting SF-36 Physical and Mental Component Summary (PCS and MCS) scores for asymptomatic patients

**Figure 5.**
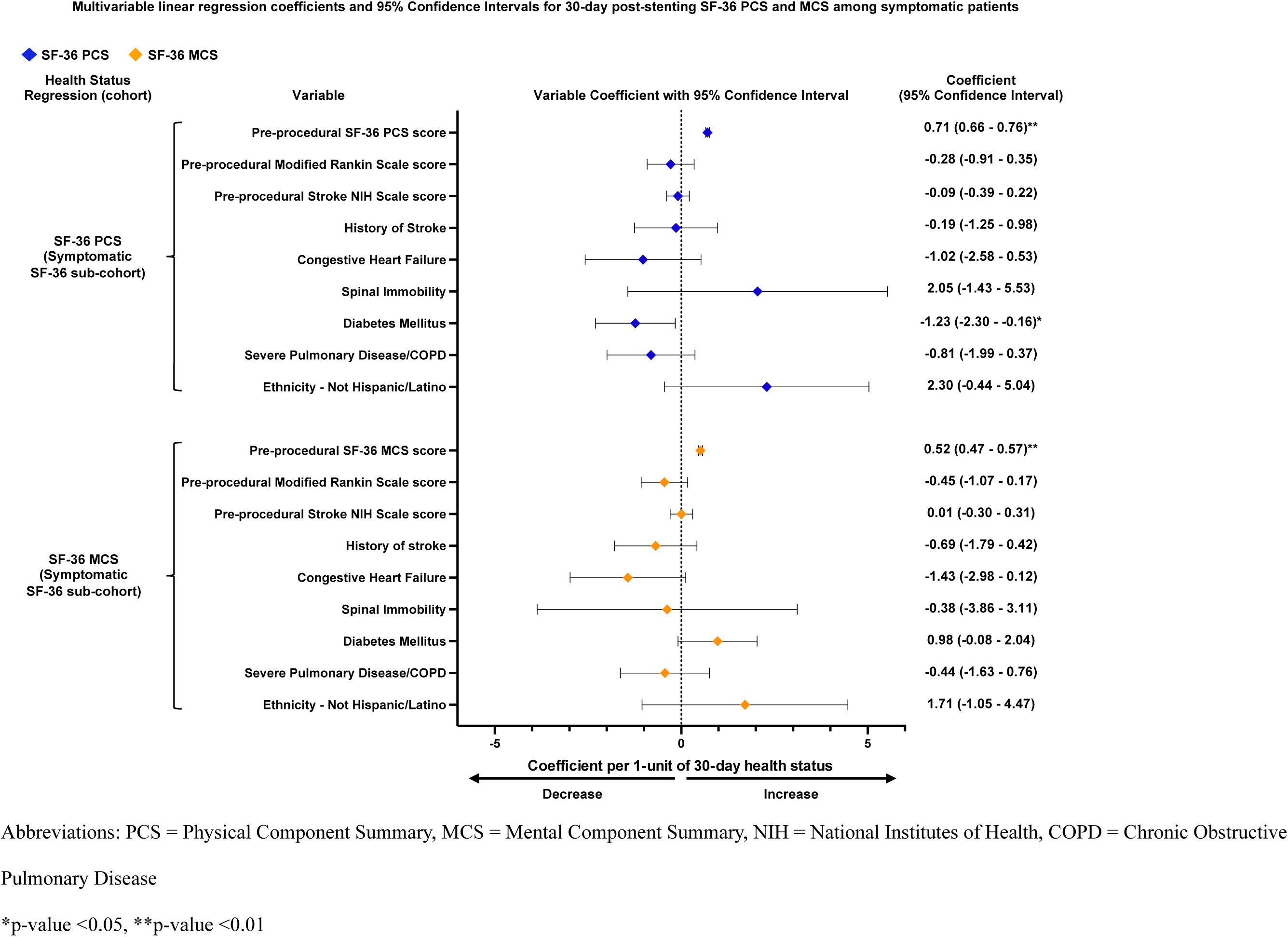
Coefficients and 95% Confidence intervals derived from multivariable linear regression models to predict 30 day post- stenting SF-36 Physical and Mental Component Summary (PCS and MCS) scores for symptomatic patients

## DISCUSSION

Our study used machine learning to develop 30-day health status prediction models for patients undergoing transfemoral-CAS. Of 66 pre-procedural variables, nine variables were identified through RF modeling as being the most important variables which capture patients’ pre-procedural health status, stroke scale scores, and medical comorbidities, our models explained ∼36%-61% of the variance in health status 30 days post-transfemoral-CAS, with SF- 36 PCS models performing best.

Health status evaluation in patients with carotid artery stenosis is emergent, with a relative lack of understanding of which patients will experience significant benefit from revascularization and what associated clinical characteristics are. Currently, no disease-specific instruments or prediction models effectively and prospectively capture multi-dimensional demographic, socioeconomic, clinical, and psychosocial determinants of patients’ health status outcomes post-revascularization. With clinical trials underway that compare treatment modalities for carotid artery stenosis, such as the ongoing Carotid Revascularization Endarterectomy vs. Stenting Trial (CREST-2), there is an urgent need to consider shared decision-making approaches in deciding which treatment modality fits best with one’s expected risk profile, and health status outcomes become invaluable in weighing those decisions.^7^ Previously reported categorization of CAS patients’ post-stenting health status changes against calculated minimal clinically important differences shows that not all carotid artery stenosis patients experience meaningful improvements in health status post-CAS, further highlighting the importance of evaluating risks vs. benefits of CAS for patients based on their characteristics.^27^ Informed shared decision-making discussions regarding treatment are crucial, especially with CMS expanding reimbursement for CAS and no longer requiring facility approval before offering CAS for carotid artery stenosis.^4,28^ Our findings will help interventionalists discuss CAS by providing insight into whether a patient can expect to see health status improvements post-stenting based on their characteristics and comorbidities. By using available information regarding a patient’s clinical picture to understand how likely a given patient is to benefit from, or be harmed by, CAS with regards to perceived quality of life given non-negligible risk of peri-procedural stroke, interventionalists and patients can better weigh risks and benefits of the procedure to make better treatment decisions.^2,4^

The nine variables we identified from the SAPPHIRE registry can reliably predict carotid artery stenosis patients’ health status changes as measured by the SF-36 PCS and MCS, explaining 61% and 54% of changes in asymptomatic and symptomatic patients’ SF-36 PCS scores post-stenting, respectively. These variables, easily obtainable through patient charts or clinical evaluation, include demographics, comorbidities, pre-procedural stroke scale scores, and pre-procedural health status. While our study used RF models, their complexity and computational demands likely limit their clinical use in the near-future. As a more feasible alternative, our nine-variable multivariable regression models may be incorporated into clinical practice to better predict physical and mental health status outcomes post-CAS as part of carotid artery stenosis treatment decision discussions. However, the SAPPHIRE registry lacks detailed information regarding patients’ psychosocial determinants of health, socioeconomic status, and cognitive status beyond gross impairment. Therefore, variables capturing these multi- dimensional aspects of health were not incorporated into our study, but may have significant utility in predicting how patients’ quality of life will change post-stenting.

Generic health status instruments to assess carotid artery stenosis patients’ health status have their limitations and may have not captured elements that are important to the disease experience. We need to develop more robust and specific health status instruments for individuals with carotid artery stenosis who contemplate invasive treatments. RF and linear regression models performed better in predicting post-stenting SF-36 PCS and MCS scores than they did in predicting EQ-5D Index Value and VAS scores, indicating that the SF-36 may correlate more strongly with commonly captured clinical characteristics among carotid artery stenosis patients than the EQ-5D. Alternatively, EQ-5D scores may be more difficult to predict or model, as indicated by prior research.^29^ Regardless, existing health status instruments are not enough. We need to develop carotid artery stenosis-specific health status instruments which capture all relevant aspects of health status for these patients to develop more accurate health status prediction models and enhance shared decision-making when choosing whether or not to undergo CAS. Until better instruments are developed, the SF-36 may be a more appropriate clinical health status instrument for prediction based on clinically available characteristics, than the EQ-5D for carotid artery stenosis patients.

Our study should be validated and expanded upon. The development of new, more comprehensive carotid artery stenosis patient registries which include more detailed information regarding patients’ clinical characteristics, psychosocial determinants of health, treatment preferences, and socioeconomic status is an important future direction. Such registries would improve our ability to discern which patient characteristics are most predictive of patients’ health status changes post-revascularization. Future work should also examine health status changes for patients undergoing alternative revascularization procedures, such as CEA or transcarotid artery revascularization. While we chose a random forest-based approach, future studies should validate whether alternative statistical models yield improved predictive power in predicting patient health status post-stenting.

Our study has limitations. First, our analysis may not be generalizable to patients undergoing revascularization procedures other than transfemoral-CAS or to patients outside of the SAPPHIRE Worldwide registry. Second, the scope of patient information captured by the SAPPHIRE Worldwide Registry is limited and the inclusion of more detailed patient information may have altered our results. Third, we evaluated health changes based on health status scores 30 days post-stenting, which may not capture changes to patients’ health status >30 days post-stenting. Future work should evaluate patient health status changes at further time intervals post-stenting.

Using a machine learning-based approach, we developed the first predictive linear regression models for health status variation 30 days post-transfemoral-CAS for carotid artery stenosis patients. Our model encapsulated information regarding patients’ pre-procedural health status, stroke scale scores, and medical comorbidities, and may help guide interventionalists’ decision-making when considering transfemoral-CAS for asymptomatic and symptomatic carotid artery stenosis patients.

## Data Availability

Data for this manuscript was obtained from the SAPPHIRE Worldwide Registry

## ACKNOWLEDGEMENTS

SAPPHIRE Worldwide investigators and contributors.

